# Brain-SAM: A SAM-based Model Tailored for Brain MRI Lesion Segmentation

**DOI:** 10.64898/2026.01.30.26345164

**Authors:** Yinghong Pan, Xiaotong Yuan, Honglei Liu, Yuqing Yang, Guixia Kang

**Affiliations:** Beijing University of Posts and Telecommunications; Capital Medical University

## Abstract

Magnetic resonance imaging (MRI) is a cornerstone of modern neuroimaging, where accurate segmentation of brain structures and lesions is essential for diagnosis, treatment planning, and clinical research. However, most current foundation models are trained on mixed-organ datasets, while the anatomical structures of the brain differ substantially from those of other organs such as the lungs and kidneys. As a result, these models often struggle to adapt to the distinctive characteristics of brain tissue. In this work, we present Brain-SAM, a model tailored for brain MRI segmentation. Brain-SAM extends the Segment Anything Model 2 (SAM2) framework by enabling the Hiera encoder to directly process 3D volumetric data and introducing a UNETR-inspired decoder for hierarchical feature decoding. The model preserves the interactive segmentation paradigm of SAM while also supporting fully automatic segmentation. Trained on multiple brain MRI datasets covering brain tumors, stroke, and epilepsy, Brain-SAM demonstrated superior performance to state-of-the-art methods. Compared with nnU-Net, it achieved Dice scores improvements of 22%, 9%, and 6% on epileptic lesions, brain metastases, and meningiomas, respectively. Notably, Brain-SAM showed clear advantages in small-lesion segmentation, achieving 15%-18% higher Dice compared with other strong baseline models. We believe that Brain-SAM may offer a useful pre-trained model for downstream brain MRI analysis tasks, and could contribute to future research and clinical applications.Our code and models are available at https://github.com/DLbrainsam/Brain-SAM.

## 1 Introduction

Magnetic resonance imaging (MRI) has become a cornerstone of modern neuroimaging, offering high-resolution, non-invasive visualization of brain anatomy and pathology[1]. It plays a critical role in the diagnosis, treatment planning, and longitudinal monitoring of neurological conditions such as tumor, stroke, epilepsy, and neurodegenerative disorder[2]. Accurate segmentation of brain MRI is indispensable for quantifying tissue structures, delineating lesions, and supporting both clinical decision-making and large-scale population studies.

Early approaches to brain MRI segmentation relied on traditional image processing and statistical modeling techniques, such as atlas-based registration[3, 4, 5], deformable models [6, 7, 8], and clustering algorithms[9, 10, 11, 12]. While these methods provided valuable insights, their performance was often constrained by sensitivity to noise, variations in imaging protocols, and limited ability to capture complex anatomical structures.

With the advent of deep learning, convolutional neural networks(CNNs)[13] rapidly became the dominant paradigm. Representative models such as U-Net [14] and its numerous variants[15, 16, 17, 18] achieved remarkable improvements in segmentation accuracy. However, due to the inherently local receptive fields of convolution operations, CNNs struggle to model global contextual relationships, often leading to suboptimal delineation of tumor boundaries from surrounding tissues. More recently, Transformer[19]-based models [20, 21, 22, 23] have been introduced, leveraging self-attention mechanisms to capture long-range dependencies and demonstrating performance comparable to or even surpassing that of CNNs. Nevertheless, while Transformers excel at modeling global context, they are less effective at capturing fine-grained local textures and boundary details, resulting in inferior preservation of local structural information. Moreover, both CNN- and Transformer-based methods are typically designed and trained for specific datasets or segmentation tasks, and their performance often deteriorates when transferred to new domains. This task-specific nature fundamentally limits their scalability and generalizability in real-world clinical applications.

In recent years, foundation models for segmentation have emerged, among which the Segment Anything Model (SAM) [24] and its upgraded version, SAM2[25], stand out as representative examples. Leveraging a prompt-based segmentation paradigm, SAM has demonstrated remarkable generalizability in the natural image domain, enabling cross-task transfer and zero-shot segmentation, and is thus regarded as a major milestone in the field of image segmentation. In the medical image domain, researchers have adapted SAM to various organ and lesion segmentation tasks, such as MedSAM [26], SAM-Med2D[27], SAM-Adapter[28], and SAM-Med3D[29]. These adaptations have shown promising cross-organ and cross-disease potential, effectively addressing long-standing challenges in medical image segmentation, including limited data availability, modality heterogeneity, and high annotation costs.

Despite recent advances, SAM-based medical segmentation models still face significant challenges in brain MRI analysis. Models such as MedSAM and SAM-Med3D are trained on large-scale, multi-organ datasets that include structures like the heart, liver, and lungs, as well as some brain data. However, the anatomical differences between the brain and other organs are substantial:the brain exhibits complex tissue organization, indistinct gray–white matter boundaries, and highly diverse lesion morphologies. Such mixed-organ training limits the models’ ability to capture the fine-grained structural characteristics of the brain. In addition, many medical image modalities, such as MRI, are inherently three-dimensional(3D), yet SAM was originally designed for 2D natural images. As a result, most SAM-based adaptations[26, 27] convert volumetric scans into 2D slices for frame-wise segmentation, which neglects spatial context across slices and often yields suboptimal performance. Moreover, most SAM-based approaches[26, 27, 28, 29] largely rely on prompt-based paradigms, where manual interactive inputs are required. For subtle or less apparent cases, such as epileptic lesions, clinicians with limited experience may struggle to provide accurate prompts[30]. This reliance on manual interactions constrains the feasibility of large-scale automated deployment in clinical practice. Furthermore, the mask decoder in SAM and its derivatives was originally designed for 2D natural images, where target objects typically exhibit clear and enclosed boundaries. When applied to brain MRI data—characterized by blurred edges, overlapping signals, and pronounced tissue heterogeneity—these decoders often exhibit degraded performance.

Therefore, we introduce Brain-SAM, a model tailored for MRI-based neuroimage segmentation, illustrated in Fig. 1. Our key contributions are outlined below:

**Figure 1.**
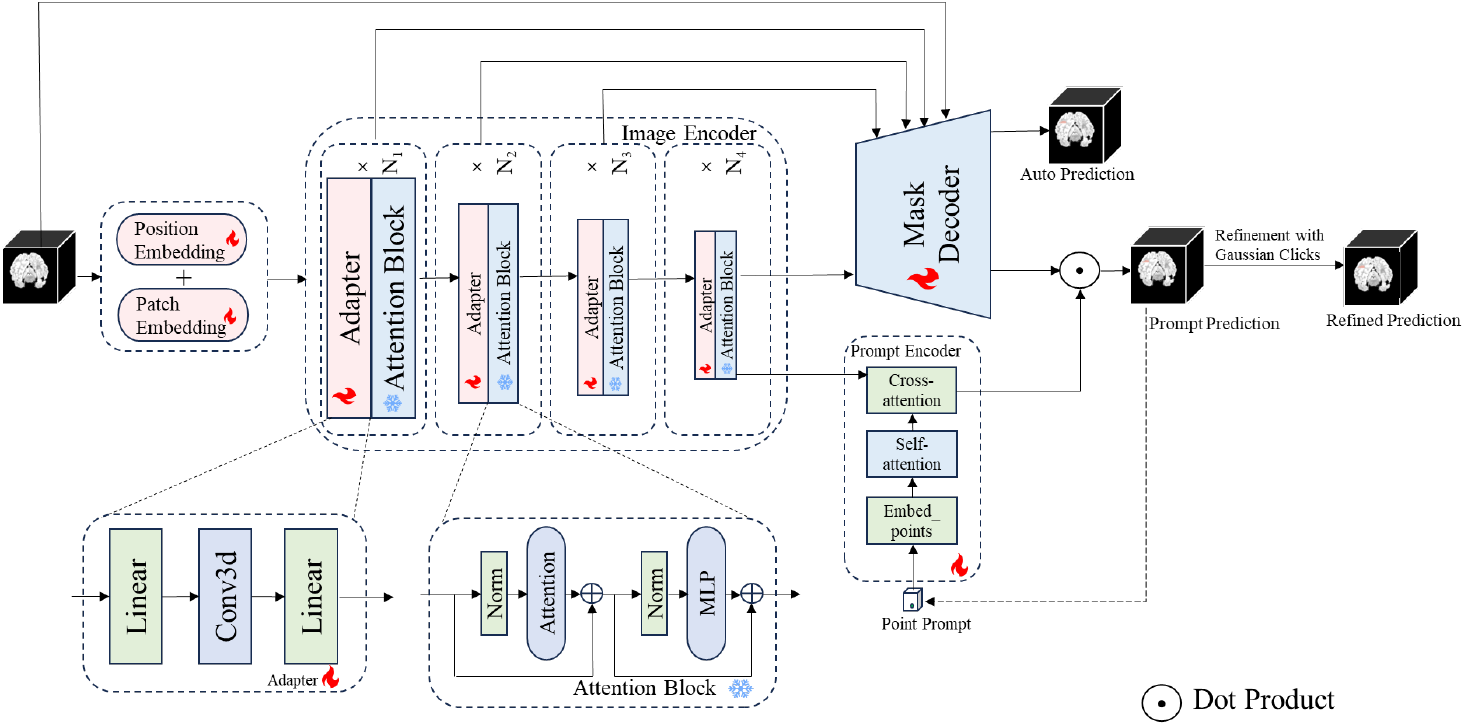
Illustration of the proposed Brain-SAM for brain lesion segmentation. The framework comprises an image encoder, a prompt encoder, and a mask decoder, supporting both automatic segmentation and prompt-based segmentation. The image encoder maps the input images into high-level features. For automatic segmentation, these features are fed into the mask decoder to generate the segmentation mask. For prompt-based segmentation, these features update the embedded prompts through cross-attention in the prompt encoder, and the updated prompts are then integrated with the second mask decoder output to produce the prompt-based prediction.

1. We extended the Segment Anything Model (SAM2) [25], which was originally developed for 2D image analysis, to 3D medical image analysis tasks. Specifically, we adapted the image encoder, prompt encoder, and mask decoder from 2D to 3D, enabling the model to directly process volumetric data rather than relying on slice-by-slice processing.
2. We introduced a UNETR[21]-inspired decoder following the SAM2 image encoder. This addition enables the model to support automatic segmentation in a fully end-to-end manner, while still retaining its original capability for interactive segmentation. In this way, Brain-SAM can handle both user-guided and fully automated segmentation workflows within a unified architecture, while also leveraging the complementary strengths of CNNs and Transformers to capture both local fine-grained details and global contextual dependencies.
3. In addition, we curated a large brain MRI dataset covering brain tumors, stroke, and epilepsy, enabling the model to support a broad range of clinical applications.

Comparative experiments on multiple publicly available datasets show that Brain-SAM achieved overall improvements over existing mainstream models in terms of both Dice score and the 95% Hausdorff distance. In addition, it exhibitted notably improved segmentation accuracy on small-target structures, such as epilepsy lesions, which are often challenging to delineate. Taken together, these improvements offer significant clinical benefits by providing more reliable and accurate segmentation, thereby supporting better-informed clinical decision-making. The code of Brain-SAM is available at https://github.com/DLbrainsam/Brain-SAM.

## 2 Method

In this study, we introduce the Brain-SAM to effectively segment lesions in MRI scans. The segmentation task is divided into two modes: automatic segmentation and prompt-guided segmentation. As illustrated in Fig. 1, the overall architecture of Brain-SAM is similar to SAM, consisting of three components: an image encoder, a prompt encoder, and a mask decoder. Automatic and prompt-guided segmentation share the image encoder and most parameters of the mask decoder, with only the final layer of the decoder employing separate weights.

### 2.1 Image Encoder

The image encoder in SAM2 is Hiera[31], which employs a hierarchical Vision Transformer (ViT)[32] architecture to efficiently capture both local and global context. Its multi-scale design enables rich feature representations at multiple resolutions, improving segmentation accuracy.

In this study, the original 2D Hiera is adapted to handle 3D medical images by extending four key modules. First, in the Patch Embedding stage,3D convolution (Conv3D) is employed to partition volumetric inputs into voxel patches. Second, the Position Embedding is extended from a 2D representation to a 3D form, enabling the model to capture spatial relationships across three dimensions. Third, although the Attention module itself is agnostic to spatial dimensionality, the number of input tokens must be expanded when moving from 2D to 3D. Specifically, a 2D feature map is flattened along its height and width to form a sequence of tokens, while a 3D volume is flattened along depth, height, and width, resulting in a longer token sequence suitable for volumetric attention. Furthermore, Q-Pooling is extended from 2D to 3D by replacing MaxPool2d with MaxPool3d, allowing feature downsampling jointly across depth and spatial dimension to support hierarchical modeling of volumetric data. With these extensions, the 2D Hiera encoder can be seamlessly adapted to 3D medical image tasks.

### 2.2 Mask Decoder

For the mask decoder, we adopt a UNETR[21]-inspired design that integrates the global modeling capacity of Transformers with the local fine-grained representation of CNNs via multi-scale feature fusion. Unlike the original UNETR decoder—designed for pure ViT encoders with uniform feature resolutions and thus requiring resolution transformations—our framework leverages the Hiera encoder, which inherently provides hierarchical features at multiple spatial scales and semantic levels. These multi-resolution features can be directly fused and progressively upsampled by the decoder to reconstruct the final segmentation output, eliminating the need for additional resolution adjustments.

Moreover, the automatic and prompt-based segmentation tasks share the same image encoder and most layers of the decoder. The two tasks differ only in the final convolutional layer, where the decoder splits into two heads: one generates the segmentation mask for automatic segmentation, while the other produces intermediate feature representations for prompt-based segmentation.Formally, let *D*_share_ denote the shared decoder backbone, while *H*_auto_ and *H*_prompt_ represent the final convolutional layers for automatic segmentation and intermediate representation, respectively. Given the image feature *F*_img_, the outputs are computed as follows:

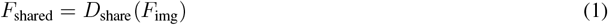

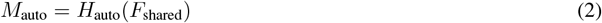

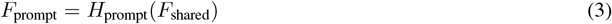

where *M*_auto_ denotes the segmentation mask predicted for the automatic segmentation task, and *F*_prompt_ represents the intermediate feature output for prompt-based segmentation.

### 2.3 Pretraining Strategy

Since the Hiera backbone in SAM2 was originally designed for 2D natural image tasks, substantial architectural modifications were introduced to adapt it for 3D medical image inputs. To obtain stable and generalizable medical representations, we initialized the model with the pretrained SAM2 image encoder weights and conducted a pretraining stage on the BraTS2021 dataset[33], fully training the model on 1000 cases across four modalities (4000 volumetric images in total). During this stage, the backbone was trained exclusively for automatic segmentation without incorporating any prompt-based mechanisms, ensuring that its representational capacity primarily stemmed from modeling medical images. The model was trained for 800 epochs until convergence was essentially achieved. Importantly, the BraTS2021 dataset was used solely for pretraining and was excluded from all subsequent training and evaluation.

### 2.4 Prompt Encoder

Among different types of prompts, bounding boxes are impractical for 3D medical images due to their complexity, while text prompts are hindered by the modality gap and difficulties in semantic alignment, especially for precise voxel localization. Therefore, point prompts are adopted as a more practical and effective choice.

The 2D prompt encoder in SAM2 is extended to a 3D version, in which all related operations, including point embeddings, are converted to three-dimensional forms to accommodate volumetric inputs. The resulting 3D point embeddings are then combined with the 3D image features extracted by the image encoder via a cross-attention mechanism, to guide prompt-based segmentation.

### 2.5 Prompt-based Segmentation

During both training and inference, we follow the interactive paradigm described in the original SAM, where prompt points are added iteratively and the next point is selected from the misclassified regions of the previously predicted mask. The point prompts are encoded into prompt embeddings, which serve as queries in the cross-attention operation with the image features (keys/values) from the image encoder. However, our mask generation strategy differs from that of SAM. In SAM, cross-attention updates both the queries and keys. The queries are first transformed by a multilayer perceptron (MLP) and then combined with the keys through a dot-product operation to produce the mask logits. In contrast, our design retains the processed queries but replaces the keys in the dot-product operation with the intermediate feature representations from the UNETR-inspired decoder,which are defined in Section 2.2. The operation can be formally expressed as:

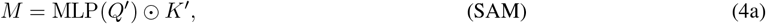

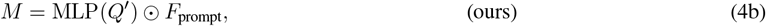

where *Q*^*′*^ and *K*^*′*^ denote the queries and keys representations updated via cross attention, ⊙ denotes the dot product operation,*F*_prompt_ corresponds to the intermediate feature representations from the UNETR-inspired decoder, and *M* corresponds to the generated mask logits.

### 2.6 Adapter-based Fine-tuning

To preserve the general representation capacity of our pretrained backbone and enable efficient adaptation to multiple brain lesion datasets, lightweight adapter modules are introduced before each block of the Hiera backbone, as illustrated in Fig. 1. Each adapter adopts a bottleneck architecture defined as

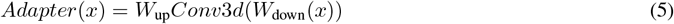

where *W*_down_ and *W*_up_ are learnable linear projections that reduce and restore the feature dimension, respectively, and *Conv*3*d*(·) denotes a 3D convolutional layer that extracts informative representations from volumetric data. During fine-tuning, only the adapter parameters are updated, while the pretrained image encoder remains frozen to prevent catastrophic forgetting of previously learned representations. Moreover, rather than maintaining separate adapters for each dataset, a shared adapter is employed across all tasks, enabling the model to learn a unified feature space that captures anatomical commonalities. This design is parameter-efficient, enables low-cost training, and supports effective cross-dataset adaptation by leveraging pretrained representations.

### 2.7 Refine

For prompt-based segmentation, a refinement mechanism in the form of post-processing is introduced. Given a mask predicted from prompts with values in [0,1] after sigmoid activation, user-provided positive (foreground) or negative (background) clicks are used to locally refine the predicted mask. Specifically, for each click, a fixed-radius 3D Gaussian kernel is centered on the click location, defining a local neighborhood around the click. The kernel is used to softly propagate the influence of the click to the neighboring voxels in the predicted mask. Positive clicks increase the mask probabilities within the local neighborhood, while negative clicks decrease them. In this way, the model can leverage click information more stably and effectively, improving both the accuracy and robustness of prompt-based segmentation.

### 2.8 Loss Function

The loss function is defined as a combination of Dice loss and Focal loss, and can be formulated as follows:

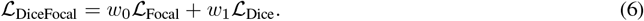

where the Dice loss is defined as:

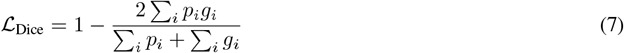

and the Focal loss is defined as:

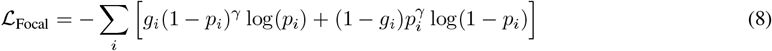

Here,*p*_*i*_ and *g*_*i*_ denote the predicted probability and ground truth label for voxel *i*, and *γ* is the weighting parameter for Focal loss. In our experiments,*γ* is set to 3 and the weights *w*_0_ and *w*_1_ are both set to 0.5. The total loss over multiple simulated user interactions is defined as:

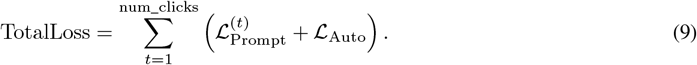

where the prompt-guided loss at interaction *t* is defined as

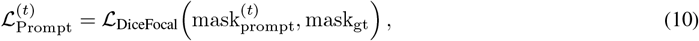

and the automatic loss is defined as

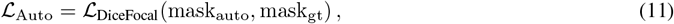

where mask_gt_ denotes the ground-truth segmentation mask, 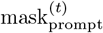 denotes the predicted segmentation mask at interaction *t* in the prompt-guided mode, and mask_auto_ denotes the predicted segmentation mask in the automatic mode.

## 3 Experiments

### 3.1 Datasets

We curated a collection of 6,768 volumetric MRI scans from publicly available datasets, including BraTS-PED [34], BraTS-SSA[35], BraTS-MET[36], BraTS-MEN[37],ISLES2022[38], ATLAS v2.0[39], and OPMRI[40]. Together, these datasets span six MRI modalities: DWI, ADC, T1w, T1ce, T2w, and FLAIR. And these scans can be categorized into three disease groups: brain tumors (BraTS-PED, BraTS-SSA, BraTS-MEN, BraTS-MET), stroke (ISLES2022, ATLAS v2.0), and epilepsy (OPMRI).

During preprocessing,non-brain tissues were removed through skull-stripping. All images were then cropped and resampled to a standardized size of 128 × 128 × 128 voxels, with an isotropic voxel spacing of 1 × 1 × 1 mm^3^.Each of the seven datasets was divided into training and test sets.

### 3.2 Competing Task-Specific and Foundation Models

To comprehensively evaluate the effectiveness of Brain-SAM, comparisons were conducted with both task-specific expert models and generalist foundation models under single-modality input settings. For the task-specific expert models, instead of training on each modality separately, all available modalities within a dataset were combined and jointly trained as a single task. For the generalist foundation models, training was performed on the combined training sets from all datasets, with modalities from different datasets also merged together. To prevent data leakage, all modalities from a subject were confined to either training or testing.

The task-specific expert models included the following:

nnU-Net[18](task-specific): Widely regarded as the state-of-the-art (SOTA) in medical image segmentation. Separate nnU-Net models were trained for each dataset, with each treated as an independent task.

UNETR[21]: A hybrid architecture with a Vision Transformer–based encoder for capturing global context and a CNN-based decoder for reconstructing the segmentation maps.

Swin UNETR[41]: A hybrid encoder–decoder architecture, where the encoder integrates Swin Transformer[42] blocks for hierarchical feature learning, while the CNN-based decoder produces the segmentation outputs.

The foundation models included the following:

nnU-Net[18] (foundation): In addition to task-specific training, a single nnU-Net was trained across all datasets, which can be regarded as a foundation model.

SAM-Med3D[29]: A SAM-based model pretrained on large-scale 3D medical images, used for prompt-based segmentation with a 6-point prompt applied during inference.

VISTA3D[43]: A generalist model based on a U-Net–type architecture, used for both automatic and prompt-based segmentation, with a 6-point prompt applied during inference.

In Brain-SAM prompt-based segmentation, a 6-point prompt is applied during inference.

### 3.3 Implementation Details

All models were trained for 300 epochs on four NVIDIA RTX A6000 GPUs (48 GB each). Brain-SAM was initialized with the weights pretrained on the BraTS2021 dataset, and during fine-tuning, the backbone of the image encoder was frozen except for the Adapter modules. SAM-Med3D and VISTA3D were initialized with the pretrained weights released by their authors. Swin UNETR and UNETR were implemented using the MONAI[44] framework. For Brain-SAM, Swin UNETR, and UNETR, training was conducted with the AdamW optimizer and a LinearLR scheduler, whereas all other baselines were trained with their default hyperparameter settings.

### 3.4 Evaluation Metrics

In the following experiments, the segmentation performance is evaluated using two complementary metrics: the Dice similarity coefficient (Dice) and the 95% Hausdorff distance (HD95).

#### 3.4.1 Dice Similarity Coefficient

The Dice is a widely adopted overlap-based metric that measures the similarity between the predicted segmentation *P* and the ground truth *G*. It is defined as:

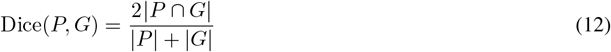

where | *P* ∩*G* | denotes the number of overlapping voxels between the prediction and the ground truth, and | *P* |, |*G* |represent the voxel counts of the predicted and ground truth masks, respectively. The Dice ranges from 0 to 1, where a value of 1 indicates perfect agreement, while 0 indicates no overlap.

#### 3.4.2 95% Hausdorff Distance

The Hausdorff distance measures the maximum distance between boundary points of the predicted segmentation and those of the ground truth. Formally, the 95% Hausdorff distance (HD95) is defined as:

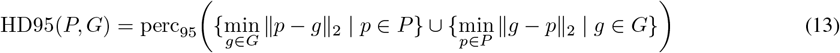

where *P* denotes the set of surface points of the predicted segmentation,*G* denotes the set of surface points of the ground truth annotation, ∥·∥_2_ represents the Euclidean distance, and perc_95_(·) denotes the 95th percentile of a given set of distances. Lower HD95 values indicate more accurate boundary alignment.

Together, Dice and HD95 provide complementary perspectives on segmentation quality: Dice evaluates volumetric overlap, while HD95 assesses boundary precision, offering a comprehensive evaluation of segmentation performance.

## 4 Result

### 4.1 Comparison with Baselines

Table 1 presents the average Dice score and HD95 (mm) of Brain-SAM compared with other models across multiple datasets. In the automatic segmentation mode (auto), our model already outperformed all foundation models in both Dice and HD95 metrics. The HD95 values were notably lower, indicating more precise boundary localization and fewer segmentation outliers, while the Dice improvements were stable and consistent. When prompts were introduced and the results were further refined, the performance improved even more. The prompt-based and refined version consistently enhanced boundary precision and overall segmentation quality across datasets, showing clear advantages over nnU-Net, SAM-Med3D, and VISTA3D. Fig. 2 presents a visual comparison between our proposed Brain-SAM and several competing methods. It can be observed that Brain-SAM exhibits a closer alignment with anatomical contours, particularly in regions where boundaries are subtle or poorly defined.

**Table 1:** Segmentation results of our method and other competing methods on public datasets.

| Category | Method | BraTS-PED |  | BraTS-SSA |  | BraTS-MET |  | BraTS-MEN |  | ATLAS v2.0 |  | ISLES2022 |  | OPMRI |  |
| --- | --- | --- | --- | --- | --- | --- | --- | --- | --- | --- | --- | --- | --- | --- | --- |
|  |  | Dice | HD95 | Dice | HD95 | Dice | HD95 | Dice | HD95 | Dice | HD95 | Dice | HD95 | Dice | HD95 |
| Task-specific Model | UNETR | 0.7561 | 5.6318 | 0.6939 | 7.4515 | 0.3964 | 18.0515 | 0.7739 | 3.7510 | 0.4297 | 14.4052 | 0.4634 | 13.9674 | 0.5334 | 6.2063 |
|  | Swin UNETR | 0.7540 | 5.5406 | 0.7203 | 6.6362 | 0.4447 | 14.1961 | 0.7881 | 3.8233 | 0.5651 | 11.5091 | 0.4769 | 12.3858 | 0.4921 | 7.3245 |
|  | nnU-Net | 0.8867 | 6.4727 | 0.8120 | 8.6478 | 0.5909 | 27.1395 | 0.7981 | 8.3652 | 0.6696 | 12.7055 | 0.6801 | 11.7763 | 0.4300 | 17.5499 |
| Generalist Model | SAM-Med3D | 0.8233 | 6.4705 | 0.7971 | 8.9532 | 0.5604 | 19.6601 | 0.7599 | 5.6755 | 0.5113 | 13.2870 | 0.5129 | 16.3379 | 0.5383 | 6.7476 |
|  | VISTA3D(auto) | 0.8685 | 2.3400 | 0.7988 | 3.8068 | 0.5769 | 14.1454 | 0.7902 | 3.3803 | 0.6166 | 9.8793 | 0.5919 | 12.7674 | 0.3826 | 9.8614 |
|  | VISTA3D(prompt) | 0.8687 | 2.4010 | 0.7976 | 3.7991 | 0.5793 | 14.2088 | 0.7906 | 3.2952 | 0.6208 | 9.7948 | 0.5914 | 12.8473 | 0.3875 | 9.8286 |
|  | nnU-Net | 0.8761 | 6.9428 | 0.8143 | 8.7242 | 0.5967 | 28.8519 | 0.7983 | 8.0370 | 0.6696 | 13.9110 | 0.6680 | 14.0478 | 0.4026 | 16.8528 |
|  | **Ours(auto)** | 0.8938 | 1.7001 | 0.8509 | 2.8716 | 0.6805 | 10.2888 | 0.8498 | 2.2307 | 0.6930 | 7.6481 | 0.6782 | 7.8648 | 0.6182 | 4.9486 |
|  | **Ours(prompt)** | 0.8950 | 1.6678 | 0.8528 | 2.8246 | 0.6844 | 10.1774 | 0.8536 | 2.1327 | 0.6959 | 7.5151 | 0.6806 | 8.0347 | 0.6241 | 4.8770 |
|  | **Ours(refined)** | **0.8952** | **1.6087** | **0.8529** | **2.6924** | **0.7157** | **7.9716** | **0.8577** | **1.9534** | **0.7082** | **6.5903** | **0.6857** | **7.1473** | **0.6396** | **4.4045** |

**Figure 2.**
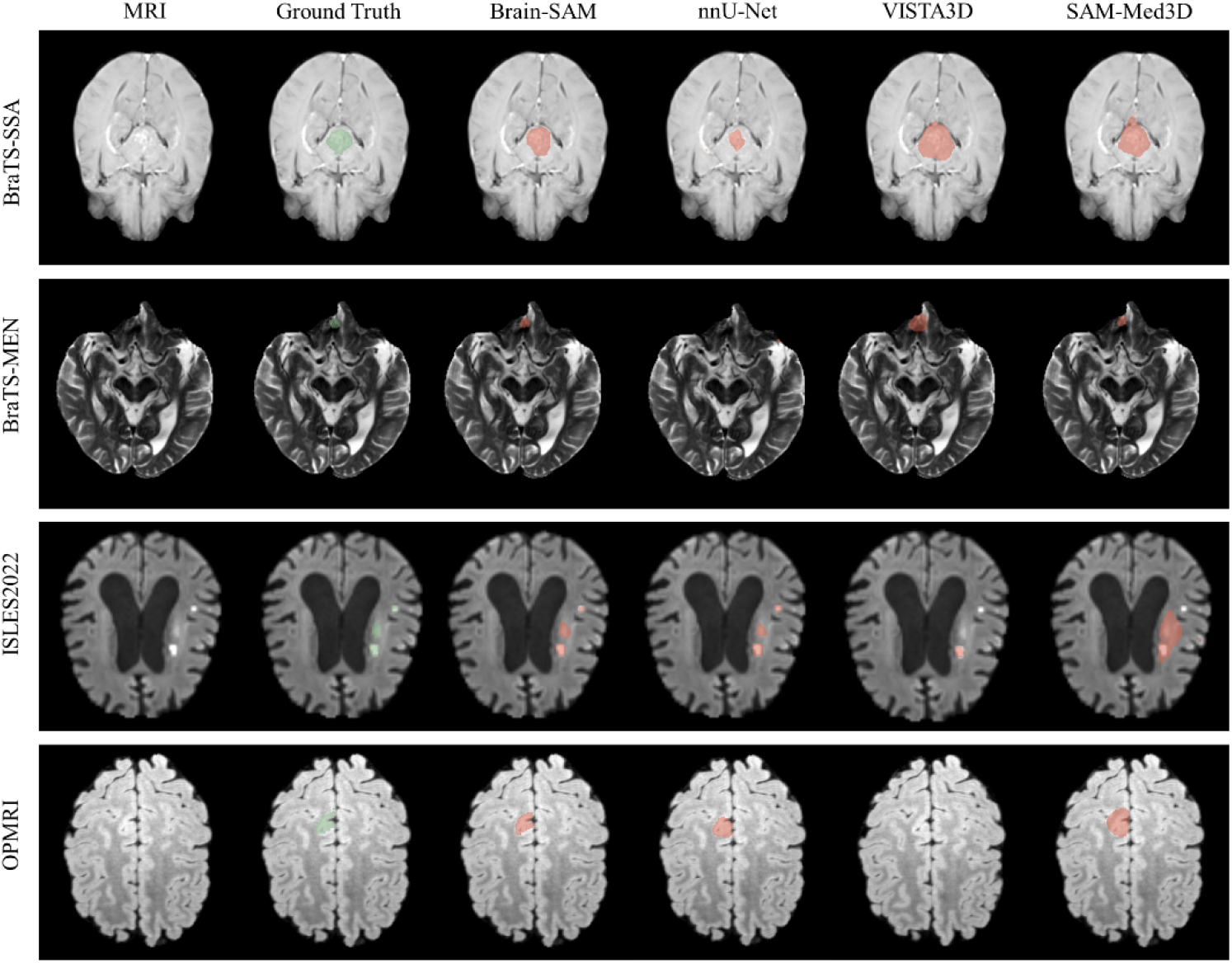
Visualization of MRI scans, ground truth, and segmentation results produced by Brain-SAM(prompt), nnU-Net, VISTA3D(prompt), and SAM-Med3D across the BraTS-SSA, BraTS-MEN, ISLES2022, and OPMRI datasets.

When analyzing performance by disease type, our model demonstrated strong results across all categories, as shown in Table 2. For the epileptic lesion dataset (OPMRI), where lesions are typically small, our approach demonstrated superior performance compared with the baseline models, achieving 8%–24% higher Dice and substantially lower HD95 than nnU-Net, VISTA3D, and SAM-Med3D.

**Table 2:**
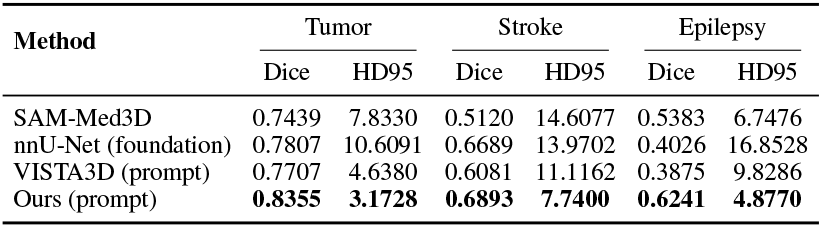
Segmentation performance of different models across brain lesion types.

| Method | Tumor |  | Stroke |  | Epilepsy |  |
| --- | --- | --- | --- | --- | --- | --- |
|  | Dice | HD95 | Dice | HD95 | Dice | HD95 |
| SAM-Med3D | 0.7439 | 7.8330 | 0.5120 | 14.6077 | 0.5383 | 6.7476 |
| nnU-Net (foundation) | 0.7807 | 10.6091 | 0.6689 | 13.9702 | 0.4026 | 16.8528 |
| VISTA3D (prompt) | 0.7707 | 4.6380 | 0.6081 | 11.1162 | 0.3875 | 9.8286 |
| **Ours (prompt)** | **0.8355** | **3.1728** | **0.6893** | **7.7400** | **0.6241** | **4.8770** |

For brain tumor (BraTS-SSA, BraTS-MEN, BraTS-PED, and BraTS-MET) and stroke datasets (ATLAS 2.0 and ISLES 2022), our model also consistently surpassed the baseline models, achieving Dice improvements of 2%–17% and notably reduced HD95 values, demonstrating robust and reliable performance across both complex tumor morphologies and irregular infarct regions.

### 4.2 Performance on Lesions of Different Sizes

The segmentation performance of different methods on lesions of varying sizes was further compared. Lesions were categorized into three groups based on the voxel count of the ground truth: small (<5,000 voxels), medium (5,000–100,000 voxels), and large (>100,000 voxels). The Dice scores of Brain-SAM(prompt), nnU-Net(foundation), VISTA3D(prompt), and SAM-Med3D across lesion size categories are summarized in the box plots shown in Fig. 3, while the corresponding recall and precision values are reported in Table 3.

**Table 3:** Performance comparison under different lesion sizes.

| Method | Large |  |  | Medium |  |  | Small |  |  | All |  |  |
| --- | --- | --- | --- | --- | --- | --- | --- | --- | --- | --- | --- | --- |
|  | Recall | Precision | F1 | Recall | Precision | F1 | Recall | Precision | F1 | Recall | Precision | F1 |
| SAM-Med3D | 0.9215 | 0.7807 | 0.8415 | 0.8330 | 0.7054 | 0.7511 | 0.7530 | 0.4265 | 0.5072 | 0.8225 | 0.6284 | 0.6880 |
| VISTA3D (prompt) | 0.8746 | 0.8941 | 0.8806 | 0.7948 | 0.8466 | 0.8081 | 0.5491 | 0.6058 | 0.5458 | 0.7350 | 0.7791 | 0.7384 |
| nnU-Net | 0.9032 | 0.8902 | 0.8943 | 0.8399 | 0.8565 | 0.8395 | 0.5407 | 0.5876 | 0.5477 | 0.7542 | 0.7755 | 0.7547 |
| Ours (prompt) | 0.9008 | 0.9079 | 0.9023 | 0.8426 | 0.8614 | 0.8446 | 0.7244 | 0.7119 | 0.6968 | 0.8144 | 0.8211 | 0.8068 |

**Figure 3.**
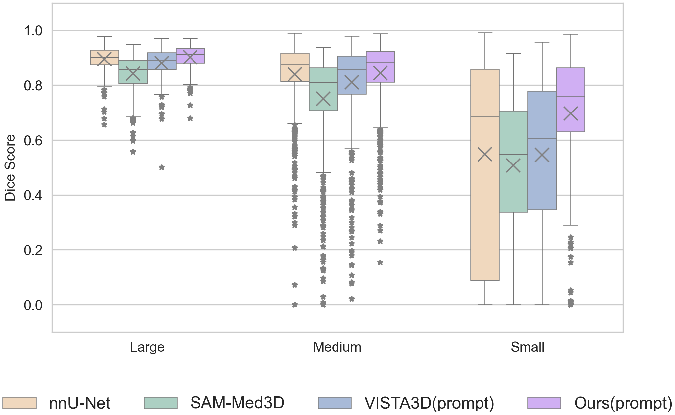
Box plots of Dice scores for Brain-SAM, nnU-Net, VISTA3D, and SAM-Med3D across lesion size categories (small, medium, and large).

As shown in Table 3, SAM-Med3D exhibited an imbalanced performance—its recall was relatively high but precision was considerably low, leading to the lowest overall F1 score. This suggested that SAM-Med3D tended to over-segment lesions and lacked boundary precision. In contrast, Brain-SAM achieved consistently higher and more balanced recall–precision pairs across all lesion sizes. For large and medium targets, Brain-SAM achieved an F1 of 0.9023 and 0.8446, respectively, slightly outperforming nnU-Net (0.8943 and 0.8395) and VISTA3D (0.8806 and 0.8081).

The advantage became most evident in small lesions, where Brain-SAM significantly surpassed the baselines, achieving higher recall (0.7244) and precision (0.7119) with the highest F1 score (0.6968), while the corresponding metrics of nnU-Net and VISTA3D were all below 0.61. These results highlighted Brain-SAM’s enhanced ability to capture subtle and fine-grained lesion details, demonstrating improved stability and robustness in small-lesion segmentation.

Moreover, the Dice score box plots in Fig. 3 further illustrate the superior performance of Brain-SAM, showing noticeably shorter boxes that indicate lower variance and thus more stable segmentation performance, with both the median and mean values being the highest among all methods. Overall, Brain-SAM consistently outperformed nnU-Net, VISTA3D, and SAM-Med3D across large, medium, and small lesions, achieving a better balance between recall and precision and demonstrating strong generalization across lesions of varying sizes.

### 4.3 Ablation Study

#### 4.3.1 Backbone Choice

To investigate the impact of different backbone architectures on segmentation performance, ablation experiments were conducted on the datasets described above, as summarized in Table 4. Consistent with SAM-Med3D, we first adopted the same mask generation strategy but replaced its image encoder from a pure 3D-ViT with 3D-Hiera. On large lesions, although 3D-Hiera achieved a slightly lower Dice score than 3D-ViT, it yielded a markedly lower HD95, suggesting more stable and accurate boundary localization. For medium and small lesions, 3D-Hiera substantially outperformed 3D-ViT, exhibiting consistently lower HD95 values. Especially on small lesions, it achieved a remarkable Dice improvement of up to 14% compared with the 3D-ViT backbone adopted in SAM-Med3D.Overall, 3D-Hiera achieved a much higher average Dice score (0.7286) than 3D-ViT in SAM-Med3D (0.6880), confirming its superior segmentation accuracy.

**Table 4:** Categories segmentation performance of different backbone architectures and mask generation strategies across lesion size categories.

| Method | Large |  | Medium |  | Small |  | All |  |
| --- | --- | --- | --- | --- | --- | --- | --- | --- |
|  | Dice | HD95 | Dice | HD95 | Dice | HD95 | Dice | HD95 |
| SAM-Med3D | 0.8415 | 7.8415 | 0.7512 | 9.0913 | 0.5073 | 9.4328 | 0.6880 | 9.9852 |
| Ours w/o UNETR Decoder | 0.8138 | 3.7286 | 0.7539 | 5.7968 | 0.6433 | 6.2938 | 0.7286 | 5.2938 |
| Ours with UNETR Decoder | 0.9023 | 1.5260 | 0.8446 | 3.7151 | 0.6968 | 5.6478 | 0.8068 | 3.9592 |

Furthermore, by integrating a UNETR-inspired decoder with the proposed mask generation strategy, the model achieved comprehensive improvements across all lesion size categories, consistently surpassing SAM-Med3D. Specifically, the incorporation of the UNETR-inspired decoder increased the average Dice by 8% compared with the version without it, and by 12% compared with SAM-Med3D.

#### 4.3.2 Effectiveness of Adapter and Pretraining Strategy

To evaluate the contribution of the adapter and the pretraining strategy, three experimental settings were designed by varying three factors: the use of BraTS2021pretraining, the inclusion of the adapter, and the fine-tuning strategy of the image encoder backbone (frozen vs. full-parameter), as summarized in Table 5.

**Table 5:**
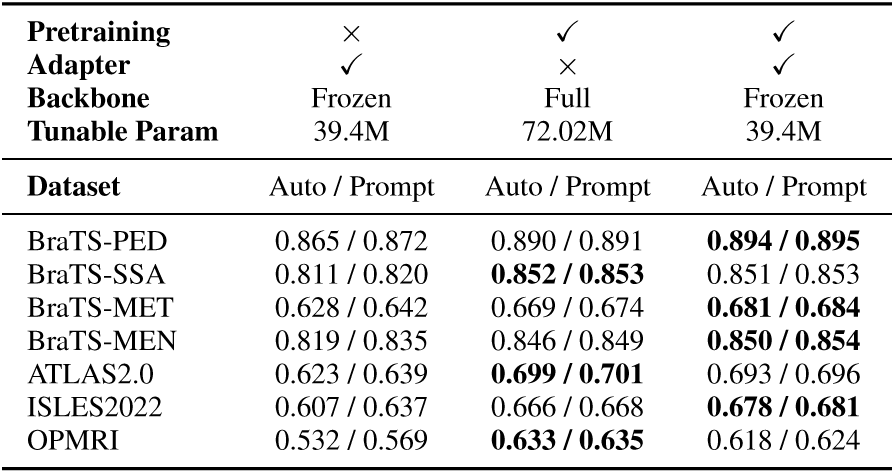
Ablation results for pretraining, adapter, and backbone fine-tuning strategies.

As shown in Table 5, using our BraTS2021-pretrained weights substantially improved segmentation performance compared to initializing with the original SAM2 weights, where the image encoder backbone was frozen and only the adapters were trained in both cases. Building upon our pretrained weights, fine-tuning only the adapters achieved nearly the same Dice score as full fine-tuning, while requiring significantly fewer trainable parameters (39.4M vs. 72.02M).

Overall, these results confirm the importance of pretraining, as the gap between natural and medical images remains substantial. The use of adapters further enhances parameter efficiency, enabling effective model adaptation with significantly fewer trainable parameters.

## 5 Discussion

The results presented in Table 1 demonstrate that Brain-SAM achieves a favorable balance between Dice score and HD95 across multiple datasets, outperforming the compared baseline models. Moreover, the stratified analysis by lesion size in Table 3 indicates that Brain-SAM maintains a good trade-off between recall and precision, with both metrics remaining relatively high—particularly for small lesions, where existing methods typically struggle, and Brain-SAM shows a remarkably superior performance.

From a clinical perspective, achieving a balanced improvement in both volumetric overlap and boundary precision is essential for brain tumor segmentation. Accurate delineation of lesion boundaries directly enables reliable volumetric assessment, which is crucial for treatment planning and longitudinal monitoring. Notably, the enhanced performance on small lesions carries significant clinical value, as subtle or early-stage abnormalities such as epilepsy-related lesions often exhibit low contrast and ambiguous margins on MRI, making them easily overlooked, particularly by less experienced clinicians[30]. The ability of Brain-SAM to maintain relatively high recall without substantially affecting precision suggests that it may serve as a supportive tool to help identify potential regions of interest and reduce the likelihood of overlooked lesions in challenging clinical scenarios.

Based on the results of our ablation experiments (Table 4), the superior performance of our Brain-SAM can be largely attributed to the image encoder and UNETR-inspired decoder.

First, regarding the image encoder, we replaced the plain ViT with the Hiera backbone. Unlike the plain ViT, Hiera performs progressive downsampling through four stages, gradually reducing the feature map resolution from 64×64×64 to 8×8×8, whereas the ViT in SAM-Med3D downsamples directly to 8×8×8 in a single step. This hierarchical design allows the model to preserve structural fidelity in the early stages, thereby retaining more fine-grained information. As a result, Hiera achieved higher Dice scores for small lesions and effectively reduced HD95 values. However, for large lesions, the performance slightly declined when using only the original SAM mask generation strategy. This is likely related to the difference in patch size between the two architectures. In our model, 3D-Hiera adopts a smaller patch size (3×3×3) compared with the 16×16×16 patches used in the 3D-ViT backbone of SAM-Med3D. For large lesions spanning broader spatial regions, the smaller patch size may limit the receptive field and weaken global context modeling, while the larger patches in 3D-ViT provide stronger long-range dependency capture, which is essential for complete lesion coverage. Future work will further investigate the influence of patch size on segmentation performance. Overall, the Hiera encoder achieved a well-balanced performance, maintaining high Dice scores while keeping HD95 low across different lesion sizes, with particularly strong performance on small lesions.

Second, to address the shortcomings of the original SAM mask decoder, we introduced a UNETR-inspired decoder. The original SAM decoder is lightweight and efficient, relying primarily on cross-attention between prompt embeddings and image features. However, due to its shallow structure, lack of multi-scale feature fusion, and limited enhancement of local details, its segmentation performance is suboptimal for small lesions and complex boundaries. In contrast, our UNETR-inspired decoder employs progressive upsampling and skip connections from the image encoder to gradually restore spatial resolution and recover high-frequency structural details, thus improving segmentation accuracy. Its output for the prompt-guided mode is fused with the prompt features to generate the final mask, thereby further enhancing the consistency and robustness of segmentation.

Overall, this hybrid design provides dual benefits: Hiera preserves fine-grained spatial details, which improves the segmentation of small lesions, while the UNETR-inspired decoder, through multi-level feature fusion, mitigates the fragmented spatial representation caused by small patch embeddings and enhances contour integrity for large lesions. Together, these complementary components maintain high Dice scores and low HD95 across different lesion sizes, resulting in a balanced and robust segmentation performance.

Nevertheless, our current study is limited to single-modality MRI data. While this setting provides a controlled environment to validate the effectiveness of our framework, it may not fully exploit complementary information across modalities. In future work, we plan to extend our approach to multi-modality integration (e.g., combining different MRI sequences), which is expected to further enhance lesion characterization and improve the robustness of segmentation across diverse clinical scenarios.

## 6 Conclusion

This paper introduces Brain-SAM, a model specifically designed for brain lesion segmentation. Brain-SAM integrates Hiera’s hierarchical encoder with a UNETR-inspired decoder while retaining the efficient prompt-interaction mechanism of SAM, enabling both automatic and prompt-guided segmentation. Extensive experiments on seven public datasets demonstrated that Brain-SAM achieved a superior balance between Dice and HD95 compared to competitive baseline models, with particularly strong performance across lesions of various sizes, especially small ones. By providing more accurate and reliable lesion delineation, Brain-SAM can help clinicians in early diagnosis, reduce manual annotation workload, and facilitate the development of downstream brain MRI analysis tasks.

## Data Availability

All datasets used in this study are publicly available and were openly accessible prior to the initiation of this study.
Specifically, the datasets used in this study include:
BraTS (Brain Tumor Segmentation) dataset, available at: https://www.med.upenn.edu/cbica/brats/
ATLAS (Anatomical Tracings of Lesions After Stroke) dataset, available at: https://fcon_1000.projects.nitrc.org/indi/retro/atlas.html
ISLES (Ischemic Stroke Lesion Segmentation) dataset, available at: https://www.isles-challenge.org/
OPMRI(An open presurgery MRI dataset of people with epilepsy and focal cortical dysplasia type II),available at:https://openneuro.org/datasets/ds004199

https://fcon_1000.projects.nitrc.org/indi/retro/atlas.html

https://www.isles-challenge.org/

https://openneuro.org/datasets/ds004199

